# Development of a novel healthcare discrimination measure: PreDict

**DOI:** 10.1101/2023.10.16.23291749

**Authors:** Carol R. Oladele, Rosana Gonzalez-Colaso, Arian Schulze, Tara Rizzo, The PreDict Writing Group, Marcella Nunez-Smith

**Author notes:** Corresponding Author: Carol R. Oladele, PhD, MPH Equity Research and Innovation Center at Yale School of Medicine, PO Box 208093 New Haven, CT 06520.

## Abstract

**Introduction:** Patient reported quality of care measures are widely recognized tools for healthcare system performance assessment. Yet, there are few existing patient reported quality of care measures regarding health equity, and none to specifically collect patient experiences of discrimination in health care.

**Objective:** To develop an item pool to measure patient experiences of healthcare discrimination-the Patient-Reported Experiences of Discrimination in Care Tool (PreDict).

**Methods:** Utilizing a multistage, exploratory sequential mixed methods study design, we conducted qualitative interviews (n=73) and expert panel consensus analysis to develop items to capture patient experiences of discrimination. This process plus systematic literature review identified extant items and informed *de novo* items for inclusion in the item pool. Items were developed in English and Spanish and were not represented by extant items. Following identification of the initial item pool (n=125), candidate items underwent cognitive interview testing with English (n=113) and Spanish (n=70) speaking participants to evaluate items for clarity and comprehensiveness. English and Spanish items were also evaluated by a bilingual expert panel to recommend pool items for inpatient field testing.

**Results:** One hundred and three items underwent cognitive interview testing and fifty-nine items were retained. Lack of clarity was the most cited factor for removal or revision of items. Expert panel review resulted in the removal of one additional item and the revision of seven items.

Fifty-eight candidate items were retained for inclusion in field testing and future analyses using item response theory modeling.

**Conclusion:** PreDict fills an important gap in measurement of discrimination, which is known to influence patient health outcomes. Development and testing to date demonstrate evidence of validity in characterizing the complex phenomenon of healthcare discrimination.

## Introduction

The Patient-Reported Experiences of Discrimination in Care Tool (PreDict) is a portfolio of measures that generates meaningful data that can be used in efforts to remedy persistent healthcare inequities. PreDict measures operationalize healthcare discrimination as patient-reported experiences and capture the equity domain of healthcare quality. The PreDict measures’ dynamic item library forms the foundation for future development of equity-focused quality measures. Among patient-reported measures, there are few that foreground health equity. We fill this critical measurement gap with the development of a hospital-level measurement approach for discrimination in inpatient settings, called PreDict-Inpatient. Existing patient-reported measures such as the Hospital Consumer Assessment of Healthcare Providers and Systems (HCAHPS) (1), Cultural Competency Assessment Tool (CCATH) for Hospitals (2), and the Communication Assessment Tool (CAT) capture the patient-centered domain of quality (3), but do not specifically address discrimination.

Distinct from most patient-reported healthcare quality measures, PreDict-Inpatient addresses the equity domain of healthcare quality by focusing on the construct of discrimination. A growing body of literature confirms that discrimination in healthcare is pervasive, with detrimental long-term mental and physical health effects. We know that patients who experience discrimination in healthcare exit care, receive poorer quality care, and have worse outcomes (4–7). Landmark reports such as the National Academy of Medicine’s (NAM) (formerly Institute of Medicine) *Unequal Treatment* (8), the annual Agency for Healthcare Research and Quality’s *Healthcare Disparities Report* (9), and numerous empirical studies identify healthcare discrimination as contributing to persistent racial/ethnic and socioeconomic disparities in health (7). Though racial/ethnic discrimination is the most studied, evidence show other forms of discrimination are also pervasive (e.g., English proficiency, sexual orientation). Yet, inconsistent measurement approaches that are not specific to healthcare settings limit the ability to aggregate data to identify target areas for intervention. PreDict-Inpatient is a unique measurement approach for healthcare system-level assessment of performance in the critical area of healthcare discrimination, as well as a new area of benchmarking.

Efforts to improve quality in US healthcare have generated national initiatives and legislation. The development and use of measurement approaches to quantify various dimensions of healthcare quality was mandated by the 2010 Affordable Care Act. Implementation of quality measurement programs by the Centers for Medicare and Medicaid Services (CMS) and public reporting have resulted in improved quality of care at hospitals across the nation (10). Despite overall improvements in quality indicators such as readmission, mortality, patient satisfaction, and other health outcomes, measures to profile healthcare equity have lagged (10). The National Quality Forum, which evaluates and endorses standardized quality measures, has called for the development of quality metrics that specifically foreground healthcare disparities among racial and ethnic populations and for which no current performance measure exists (11). The NAM has also highlighted the need for patient-reported quality measures to address healthcare inequities, the root causes of healthcare disparities. National organizations like the National Quality Forum have called for the creation of disparities-sentinel (i.e., disparities-specific) measures to improve the quality of healthcare delivery system performance; nevertheless, few metrics are designed to evaluate patient care experiences with the explicit aim of reducing or eliminating healthcare inequities.

Accordingly, the overarching goals of the PreDict measures are to: 1) standardize healthcare discrimination measurement, 2) reduce attributional ambiguity, 3) focus on healthcare organizations as the unit of analysis, 4) capture varied domains of healthcare discrimination, 5) have relevance across disparities populations (e.g. gender, race, socioeconomic status, religion), and 6) establish a measurement approach that recognizes the need for valid measures to monitor disparities among patients with limited English proficiency. This article describes the item development and refinement process to create an item pool for PreDict-Inpatient, a measurement approach for healthcare discrimination in inpatient settings.

## Methods

### Research Design Overview

Our conceptual framework was grounded in multiple theoretical domains, including aversive, symbolic, and modern racism theories (12–15). We used a modified exploratory, sequential, mixed-methods design to develop the PreDict item pool (16). Qualitative components utilized an emergent strategy to understand the phenomenon of healthcare discrimination. Quantitative components involved quantitative testing and validation of items to determine the PreDict measures item library. The item library was used to develop PreDict-Inpatient through a rigorous testing process outlined in the sections that follow. **Figure 1** illustrates the PreDict development process.

**FIGURE 1.**
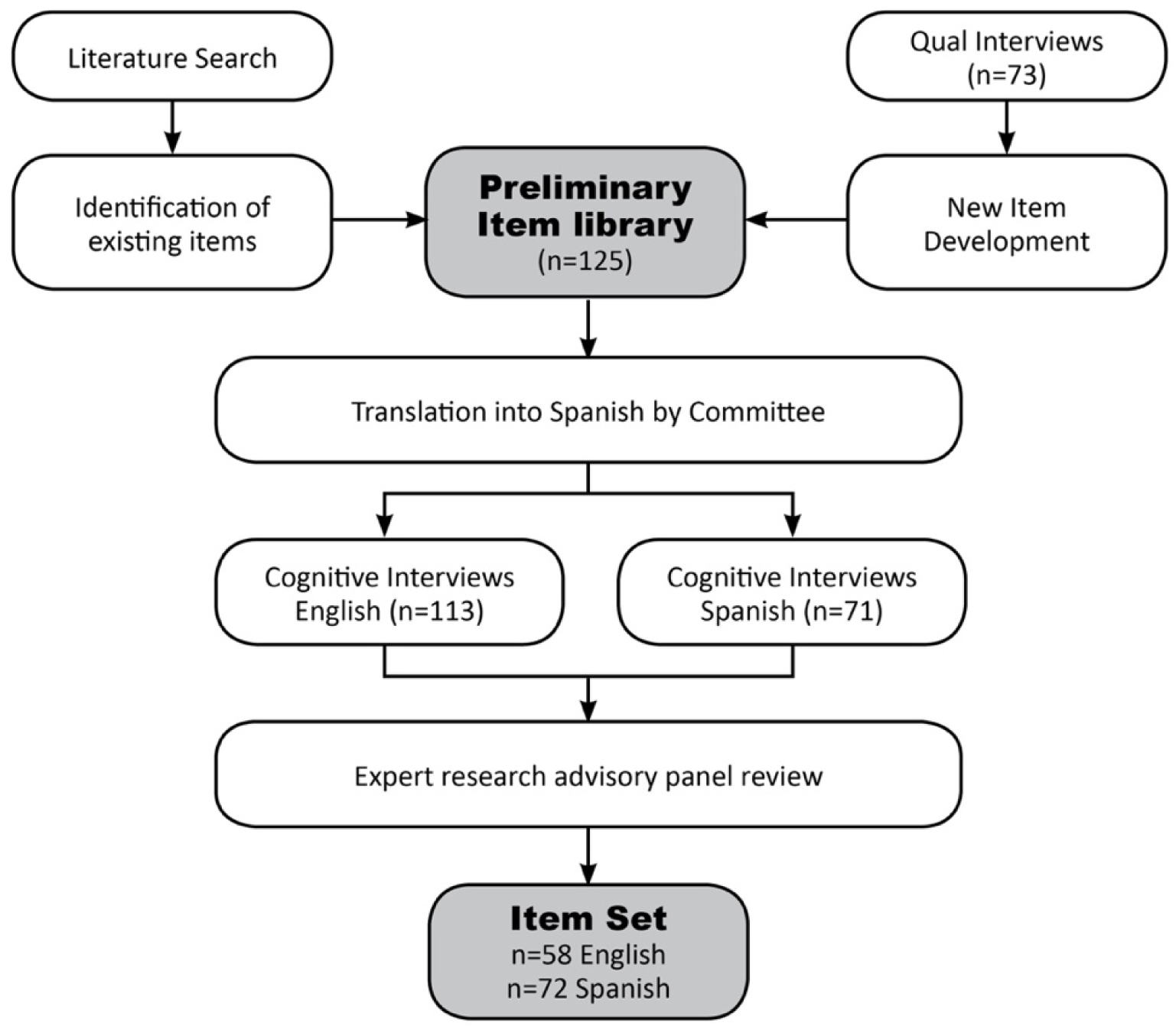
Item pool development process using qualitative methods design

**FIGURE 2.**
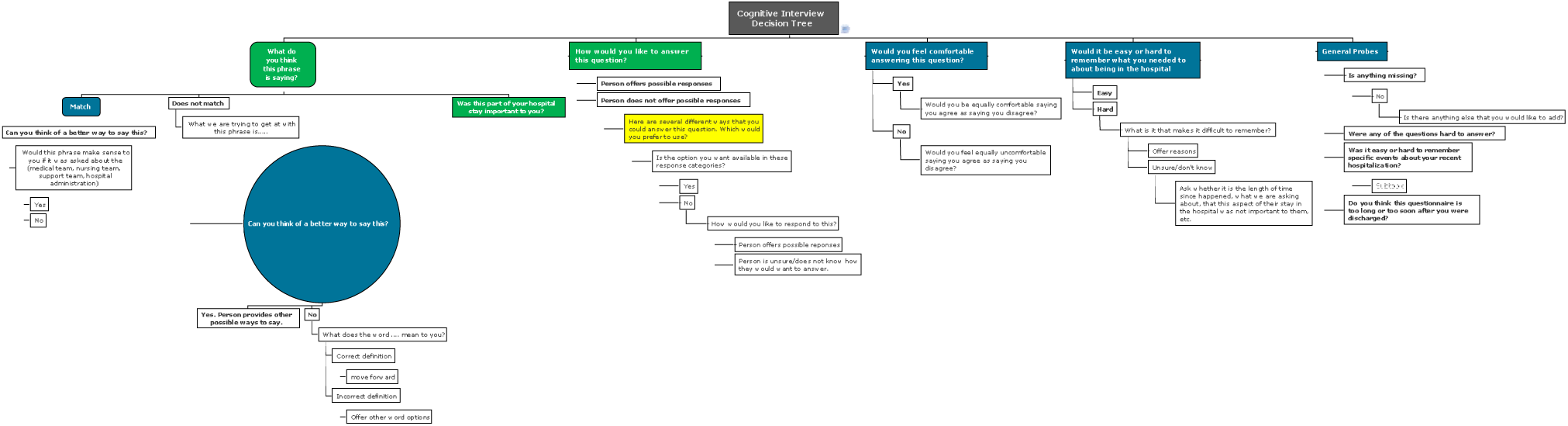
CI Decision tree

The study was approved by the Yale University Human Investigation Committee prior to data collection.

### Taxonomy

We utilized key informant interviews and reviewed extant literature to understand healthcare discrimination experiences. Domains and dimensions that emerged from analysis of qualitative interviews and extant literature review were formalized as the PreDict taxonomy to capture the latent construct of healthcare discrimination and serve as a guiding framework for item creation (17, 18). Key informant interviews were conducted among a racially and linguistically diverse sample of patients, patient advocates, healthcare providers, and payers to obtain diverse experiences of discrimination in healthcare settings. The taxonomy domains that emerged from this process included equity, information sharing, consumer service, and care effectiveness.

### Literature Searches

Literature searches were conducted to identify current quality improvement measures and avoid item redundancy in *de novo* item development. We conducted an extensive literature review on the topics of discrimination in healthcare, health inequities, and health outcomes using twenty-three search terms developed in consultation with medical librarians at Yale School of Medicine. We searched the National Library of Medicine’s MEDLINE database and the EMBASE database to capture relevant publications. We also identified and reviewed existing hospital quality measures, patient satisfaction measures, and societal discrimination measures. Relevant scales included: Communication Assessment Tool (CAT) (19), Everyday Discrimination Scale (20), HCAHPS (1), Health Care System Distrust Scale (21, 22), and Short Assessment of Health Literacy—Spanish and English (SAHL-S&E) (23). Our literature search results broadened the conceptual grounding of healthcare discrimination to inform *de novo* item development. The sections that follow detail the development methodology to create the PreDict-Inpatient item pool.

### Bilingual De Novo Item Development

A decentering approach was used to develop bilingual items for inclusion on PreDict-Inpatient. Concurrent dual language focus was critical to the development of PreDict-Inpatient as a valid bilingual measurement approach. Decentering ensures that the resulting measure is culturally and linguistically relevant and that each item is conceptually equivalent in both languages (24). Our linguistic approach aims to prevent measurement bias in conducting cross-population health research. Other measures were developed in a target language representing the linguistic majority and were then translated to other languages. The taxonomy of healthcare discrimination experiences, ratified by the research team and content expert advisors, served as the guiding framework for the creation of *de novo* items.

### Cognitive Interviews

Cognitive interviews (CIs) were conducted to test the clarity and comprehension of candidate items. Recently discharged patients who spoke English or Spanish were identified at primary care clinics in Connecticut for participation in cognitive interviews. We utilized two approaches for recruitment. At most sites, the project coordinator worked with clinic staff to identify patients who met eligibility criteria and had an upcoming appointment. Clinic staff obtained consent from patients and provided names and contact information to the research team; trained bilingual research assistants then contacted interested patients to explain the study and schedule a time for an interview. A second approach involved research assistants recruiting patients in clinic waiting areas of participating primary care clinics or clinic staff referring eligible patients to research assistants in the clinic waiting area. We were purposive in our recruitment to obtain participant diversity according to age, race, language preference, and educational attainment. Interviews were scheduled to take place at the clinic or another convenient location for the patient (e.g., libraries, eateries). Research assistants provided an information sheet to participants and explained the study again before conducting the interview. After obtaining verbal consent, the researchers administered a mini-cognitive test to participants to rule out cognitive impairment. Participants chose the interview language (English or Spanish before the interview commenced. Participants were asked about the meaning and importance of each statement, alternative phrasing, and appropriateness of response options.

A total of 184 CIs were conducted, 113 in English and 71 in Spanish. During the cognitive interview, at least two participants were asked to read and respond to each candidate items. The research assistant then probed participants on ways to rephrase the item. Participants were also asked to rate the importance of each item. Each participant reviewed seven to ten items during their interview. Participant interviews were recorded to ensure accurate documentation of feedback.

### Adjudicating cognitive interview responses

The research team reviewed cognitive interview (CI) responses during internal meetings where decisions were made to remove items that did not test well, rephrase and retest items that proved incomprehensible to some participants, and retain candidate items that tested well. The research team reviewed items by domain to identify problem items and CI response patterns within each domain. Items determined to be comprehensible and clear during CI testing were retained in the item set for future field testing. For items with mixed CI results, the research team took a conservative approach to discussion and possible revision. Items were flagged for discussion if they caused difficulty in comprehension or clarity for at least one participant. Candidate items that participants deemed to be unimportant or irrelevant in the context of discrimination were also flagged.

The research team compared responses for each item to determine the need for rephrasing and further testing. Respondent demographics were evaluated in tandem to identify whether responses were patterned by demographic characteristics. The research team decided by consensus whether to rephrase items. Items that were rephrased were then tested again during CI in both English and Spanish, and the entire process was repeated until full clarity and comprehension were achieved.

### Research Advisory Panel Review

The research team established and convened a Research Advisory Panel (RAP) to provide guidance and critical feedback at various stages of the item development process. The RAP was linguistically diverse and composed of leading researchers with expertise in a variety of areas including disparities research, racial/ethnic discrimination, measure development, psychometrics, and hospital quality improvement. The RAP reviewed and evaluated candidate items retained following the CI process.

The RAP rated each item based on four criteria: relevance, actionability, measurability, and novelty. Relevance referred to how germane each item was to the domain with which it was associated and whether it captured the latent construct of discrimination. Actionability referred to the ability of each item’s results to inform improvement efforts at participating hospitals.

Measurability rated an item’s ability to identify and distinguish concrete experiences of discrimination. Novelty of an item was determined based on the item’s uniqueness from items included in other hospital measures. Using a Qualtrics™ online survey, members also ranked the candidate items in order of importance for inclusion in the item pool. RAP members were able to provide comments on each item, providing rationale for their rating decisions.

## Results

### Cognitive Interviews

**Table 1** summarizes the demographics of English and Spanish CI participants. The mean age of English and Spanish CI participants was 51 and 54 years respectively. Forty-three and 42 percent of English CI participants self-identified as Black and White, respectively. Twenty-one percent of Spanish CI participants self-identified as White and 54% as “other.” There were more male English CI participants compared to Spanish CI participants (55% vs. 27%). Fifty-four percent of English and 86% of Spanish CI participants had a high school education or below, while 60% and 72% had yearly household incomes less than $30,000, respectively.

**Table 1.** Demographic characteristics of CI participants for English and Spanish speaking respondents.

|  | English Interviews<br>n (%) | Spanish Interviews<br>n (%) | Total<br>n (%) |
| --- | --- | --- | --- |
|  | n=113 | n=71 | 184 |
| **Mean Age** | 51.3 years | 53.5 years |  |
| **Gender** |  |  |  |
| Female | 51 (45) | 52 (73) | 103 (56) |
| Male | 62 (55) | 19 (27) | 81 (44) |
| **Hispanic/Non-Hispanic\*** |  |  |  |
| Hispanic | 18 (16) | 70 (99) | 88 (48) |
| Non-Hispanic | 94 (83) | 0 (0) | 94 (51) |
| **Race/Ethnicity\*** |  |  |  |
| White | 47 (42) | 15 (21) | 62 (34) |
| Black | 49 (43) | 3 (4) | 52 (28) |
| Multi-Racial | 2 (2) | 4 (6) | 6 (3) |
| Asian/Pacific Islander | 2 (2) | 1 (1) | 3 (2) |
| American Indian/Alaska<br>Native | 1 (1) | 2 (3) | 3 (2) |
| Other | 12 (11) | 38 (54) | 50 (27) |
| **Education** |  |  |  |
| Less than HS | 18 (16) | 42 (59) | 60 (33) |
| HS/GED | 43 (38) | 19 (27) | 62 (34) |
| Some college | 28 (25) | 6 (8) | 34 (18) |
| 4-year degree or more | 24 (21) | 4 (5) | 28 (15) |
| **Income** |  |  |  |
| Less than 10,000 | 40 (35) | 39 (55) | 79 (43) |
| 10-29,999 | 28 (25) | 12 (17) | 40 (22) |
| 30-49,999 | 15 (13) | 5 (7) | 20 (11) |
| 50-74,999 | 5 (4) | 0 (0) | 5 (3) |
| 75,000+ | 11 (10) | 0 (0) | 11 (6) |
| Don't know | 14 (12) | 15 (21) | 29 (16) |
| *Missing responses<br>excluded |  |  |  |

A total of 103 items underwent initial CI testing to identify candidate items that would progress to further testing. **Table 2** provides examples of English and Spanish items that were rephrased in response to participant feedback along with the primary reason for rephrasing. Twenty-nine items were rephrased to improve clarity, while 14 items were rephrased to improve comprehension. For Spanish items, loss of meaning was the primary reason for rephrasing. In many cases, interlanguage normative equivalence was not achieved, thereby requiring items to be revised or removed. For example, the Spanish equivalent could not be achieved for items such as “The doctors treated me like a guinea pig,” which relied on an English idiom; it was thus removed. Eight items were rephrased in both English and Spanish to achieve interlanguage equivalence. Items were removed if they routinely tested poorly or were determined to be unimportant or irrelevant by participants. Forty-five items were removed from the CI process in response to participant feedback.

**Table 2.** Examples of Items Altered in Response to Participant CI Feedback.

| Reason for Rephrasing | Original Phrasing | Revised Phrasing |
| --- | --- | --- |
| Improved Clarity | Someone asked me about my comfort with reading and writing in my primary language. | Someone asked me how comfortable I am reading and writing in English. |
| Improved Clarity | Someone explained trainees (e.g., student doctors and student nurses) would be part of teams providing my care in the hospital. | Someone explained that trainees would be part of teams providing my care in the hospital (for example: residents, student doctors and student nurses). |
| Improved Comprehension | The medical team asked me what I wanted done if my heart stopped beating or I stopped breathing. | A doctor and I talked about whether I would want them to try to save me if my heart stopped (Do Not Resuscitate conversation). |
| Improved Comprehension | I knew who to call if I had questions after leaving the hospital. | Someone explained what I should do if I had questions after leaving the hospital. |
| Improved Interlanguage Equivalence | The medical team spoke to me like I was not smart enough to understand what they were saying. | The doctors talked down to me. |

### Research Advisory Panel Review

After the CI process, the RAP evaluated a total of 59 candidate items to help refine items for inclusion in subsequent field testing. RAP members were asked to rate items using a four-point Likert scale using the following criteria: relevance, actionability, measurability, and novelty. Items were listed within their respective taxonomy domain and dimension for benchmarking during the review. RAP members were also asked to rank items in order of importance for inclusion in field testing according to domain.

We calculated the mean score for each of the four criteria based on a four-point Likert response scale (1-Strongly Agree to 4-Strongly Disagree). Overall, the candidate items were rated well with a mean score of 1.58. We took a conservative approach during the review of results and flagged an item for re-examination if it had a mean score of 2.25 or greater on any of the four criteria. Items that were identified as having little relevance to the related domain were reviewed and a determination was made to remove the item or to expand the domain definition if appropriate. We also re-examined items to assess their relevance to capturing the underlying construct of discrimination. As a result of the RAP review, the research team removed one item, reworded seven items, and redefined the definitions of one domain and one dimension in order make the definitions inclusive of the item concepts. The RAP also evaluated the suitability of item response options. Extreme responses, such as “always” and “never,” were replaced with options “almost always” and “almost never.” **Table 3** provides examples of items that were retained after RAP review and included in field testing.

**Table 3.**
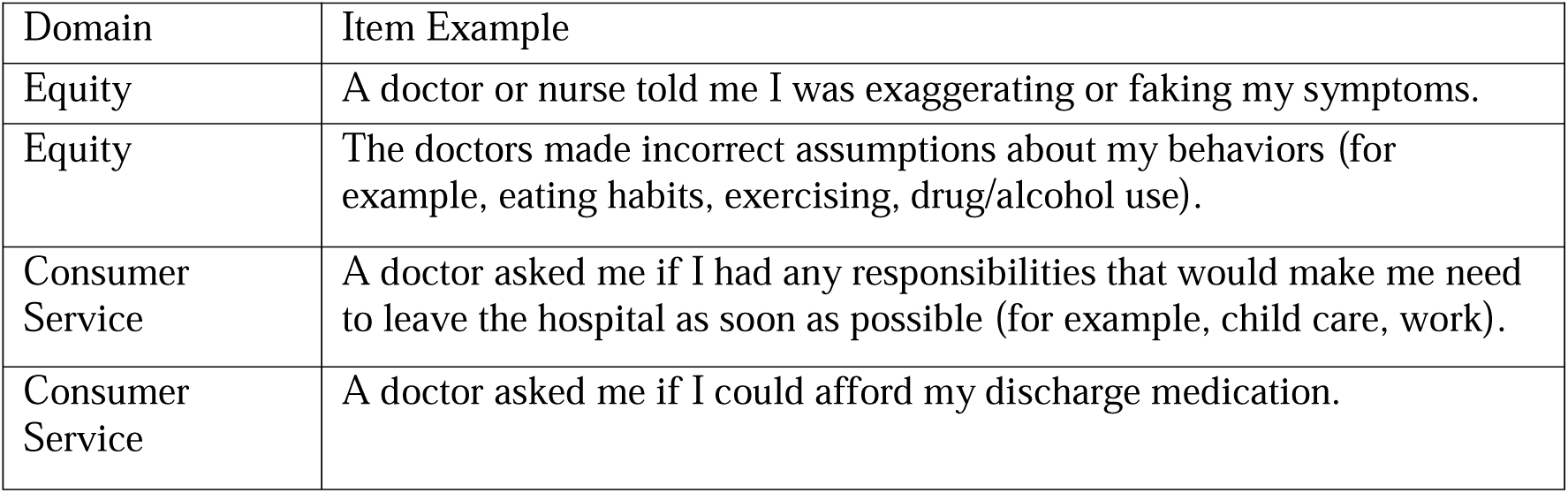
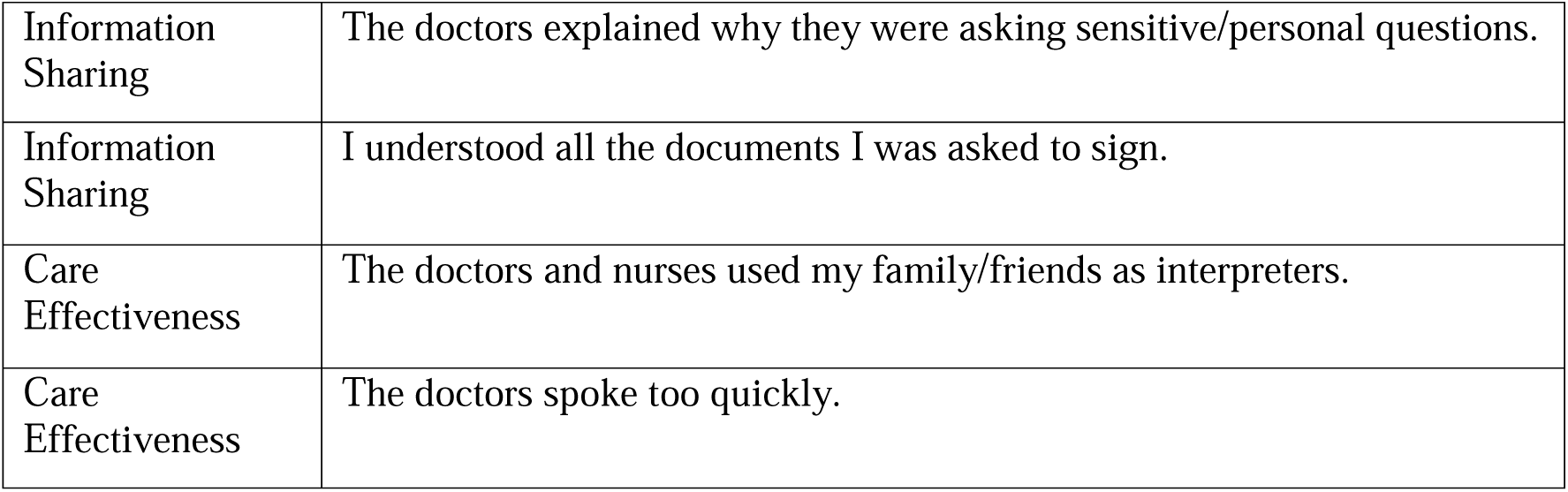
Examples of Items Retained After Expert Panel Review.

| Domain | Item Example |
| --- | --- |
| Equity | A doctor or nurse told me I was exaggerating or faking my symptoms. |
| Equity | The doctors made incorrect assumptions about my behaviors (for example, eating habits, exercising, drug/alcohol use). |
| Consumer Service | A doctor asked me if I had any responsibilities that would make me need to leave the hospital as soon as possible (for example, child care, work). |
| Consumer Service | A doctor asked me if I could afford my discharge medication. |
| Information Sharing | The doctors explained why they were asking sensitive/personal questions. |
| Information Sharing | I understood all the documents I was asked to sign. |
| Care Effectiveness | The doctors and nurses used my family/friends as interpreters. |
| Care Effectiveness | The doctors spoke too quickly. |

Fifty-eight candidate items were included in the field-testing item bank. Five hospitals across the US were then recruited for participation to test candidate items among recently discharged patients. Further testing is underway to evaluate the psychometric properties of candidate items. At the completion of field testing, we will have a final item bank of questions to measure discrimination in inpatient settings.

## Discussion

PreDict-Inpatient fills an important measurement gap that has been well noted in the healthcare discrimination literature (25). This measurement approach provides a standardized method to quantify healthcare inequities and identify areas for targeted healthcare quality improvement. Incorporating standardized measurement regarding specific discrimination domains will facilitate the development of effective health equity solutions, which is currently limited by measurement approaches that are not specific to healthcare settings, variation in measurement for healthcare discrimination, and inconsistent findings. We address limitations of current measures through application of a rigorous mixed-methods design to develop and test items that capture healthcare discrimination. The development of PreDict measures was guided by a taxonomic framework, grounded in patient perspectives, that includes multiple domains of healthcare discrimination.

This framework allowed us to develop novel items specific to healthcare settings and to utilize a multidimensional measurement approach. Additionally, our novel, non-attributional approach allows identification of other forms of discrimination besides race/ethnicity (e.g., weight, sexual orientation, gender), including emerging disparity groups. This approach facilitates examining intersectional discrimination to understand the compound effect of having multiple identities known to experience oppression and discrimination (26, 27).

Our healthcare system-based methodological approach for discrimination measurement, the first of its kind, permits the ability to evaluate healthcare system performance. Using this approach, our goal is for PreDict-Inpatient to advance healthcare equity and reduce health disparities. The observed racial and ethnic disparities related to the COVID-19 pandemic clearly demonstrate the need for disparities-specific patient reported experience measures like PreDict-Inpatient.

Innumerable tragic accounts of racial and ethnic minorities facing higher rates of healthcare access barriers, hospitalization, and mortality illuminate a critical need for measurement to understand factors that underlie disparate health outcomes.

The future adoption of disparities-specific patient experience measures like PreDict-Inpatient into existing quality measurement programs is key to identifying and monitoring disparities. Patient-reported measurements have gained increased prominence in recent years, becoming a critical component of efforts to improve quality of care and health outcomes (28–30). The creation of PreDict-Inpatient expands current domains of patient-reported outcomes to include healthcare discrimination, a phenomenon that has not been systematically studied to date. The incorporation of patient-reported outcomes measures like PreDict-Inpatient is necessary to derive solutions to address pervasive discrimination in healthcare settings. Valid and meaningful patient-reported quality measures could be instrumental in shaping policies and practices that create welcoming healthcare environments and ensure equitable care for patients. With widespread adoption and implementation, PreDict-Inpatient can facilitate progress towards improving the health of our population and narrowing gaps in health outcomes among disparity populations.

## Data Availability

All data produced in the present study are available upon reasonable request to the authors.

## References

1. Goldstein E, Farquhar M, Crofton C, et al. Measuring hospital care from the patients’ perspective: An overview of the cahps hospital survey development process. Health Serv Res. 2005;40(6 Pt 2):1977–95.

2. Weech-Maldonado R, Dreachslin JL, Brown J, et al. Cultural competency assessment tool for hospitals: Evaluating hospitals’ adherence to the culturally and linguistically appropriate services standards. Health Care Manage Rev. 2012;37(1):54–66.

3. Tzelepis F, Sanson-Fisher RW, Zucca AC, et al. Measuring the quality of patient-centered care: Why patient-reported measures are critical to reliable assessment. Patient Prefer Adherence. 2015;9:831–5.

4. Abramson CM, Hashemi M, Sánchez-Jankowski M. Perceived discrimination in u.S. Healthcare: Charting the effects of key social characteristics within and across racial groups. Prev Med Rep. 2015;2:615–21.

5. Benjamins MR, Middleton M. Perceived discrimination in medical settings and perceived quality of care: A population-based study in chicago. PLos One. 2019;14(4):e0215976.

6. D’Anna LH, Hansen M, Mull B, et al. Social discrimination and health care: A multidimensional framework of experiences among a low-income multiethnic sample. Soc Work Public Health. 2018;33(3):187–201.

7. Ben J, Cormack D, Harris R, et al. Racism and health service utilisation: A systematic review and meta-analysis. PLos One. 2017;12(12):e0189900.

8. Medicine Io. Unequal treatment: Confronting racial and ethnic disparities in health care (with cd). Washington, DC: The National Academies Press; 2003.

9. 2018 national healthcare quality and disparities report. Rockville, MD: Agency for Healthcare Research and Quality; 2018.

10. Centers for Medicare, Medicaid Services. 2015 national impact assessment of the centers for medicare and medicaid services (cms) quality measures report. Baltimore, MD: CMS. 2015.

11. Weissman J, Betancourt J, Green A, et al. Commissioned paper: Healthcare disparities measurement. Washington, DC: National Quality Forum, 2011.

12. Alderfer CP, Sims AD. Diversity in organizations. Handbook of psychology. 2003.

13. James SA. The strangest of all encounters: Racial and ethnic discrimination in us health care. Cad Saude Publica. 2017;33Suppl 1(Suppl 1):e00104416.

14. Maina IW, Belton TD, Ginzberg S, et al. A decade of studying implicit racial/ethnic bias in healthcare providers using the implicit association test. Soc Sci Med. 2018;199:219–29.

15. Williams DR, Lawrence JA, Davis BA. Racism and health: Evidence and needed research. Annu Rev Public Health. 2019;40(1):105–25.

16. Curry L, Nunez-Smith M. Mixed methods in health sciences research: A practical primer: SAGE Publications; 2014.

17. Schulze WA, Rizzo T, Oladele C, et al. Patient-reported experiences of discrimination: A taxonomy. ISMHHD 2014 International Symposium on Minority Health & Health Disparities; 2014.

18. Raskind IG, Shelton RC, Comeau DL, et al. A review of qualitative data analysis practices in health education and health behavior research. Health Educ Behav. 2018;46(1):32–9.

19. Makoul G, Krupat E, Chang CH. Measuring patient views of physician communication skills: Development and testing of the communication assessment tool. Patient Educ Couns. 2007;67(3):333–42.

20. Williams DR, Yan Y, Jackson JS, et al. Racial differences in physical and mental health: Socio-economic status, stress and discrimination. J Health Psychol. 1997;2(3):335–51.

21. Shea JA, Micco E, Dean LT, et al. Development of a revised health care system distrust scale. J Gen Intern Med. 2008;23(6):727–32.

22. Crowne DP, Marlowe D. A new scale of social desirability independent of psycholopathology. J Consult Psychol. 1960;24(4):349–54.

23. Lee S-YD, Stucky BD, Lee JY, et al. Short assessment of health literacy-spanish and english: A comparable test of health literacy for spanish and english speakers. Health Serv Res. 2010;45(4):1105–20.

24. Erkut S, Alarcon O, GarcÍA Coll C, et al. The dual-focus approach to creating bilingual measures. J Cross Cult Psychol. 1999;30(2):206–18.

25. Shavers VL, Fagan P, Jones D, et al. The state of research on racial/ethnic discrimination in the receipt of health care. Am J Public Health. 2012;102(5):953–66.

26. Crenshaw K. Demarginalizing the intersection of race and sex: A black feminist critique of antidiscrimination doctrine, feminist theory and antiracist policies. University of Chicago Legal Forum. 1989;1989(1):139–67.

27. Grollman EA. Multiple disadvantaged statuses and health:The role of multiple forms of discrimination. J Health Soc Behav. 2014;55(1):3–19.

28. Øvretveit J, Zubkoff L, Nelson EC, et al. Using patient-reported outcome measurement to improve patient care. Int J Qual Health Care. 2017;29(6):874–9.

29. Calvert M, Kyte D, Price G, et al. Maximising the impact of patient reported outcome assessment for patients and society. BMJ. 2019;364:k5267.

30. Anhang Price R, Elliott MN, Zaslavsky AM, et al. Examining the role of patient experience surveys in measuring health care quality. Med Care Res Rev. 2014;71(5):522–54.

